# Loss of SARS-CoV-2 Seropositivity among Healthy Young Adults over Seven Months

**DOI:** 10.1101/2022.07.15.22277688

**Authors:** C. Suzanne Lea, Kristina Simeonsson, Aaron Kipp, Charleen McNeill, Lisa Wilcox, William Irish, Hannah Morris, Omar Diaz, John Fallon, Rachel L. Roper

**Affiliations:** Department of Public Health, Brody School of Medicine, East Carolina University; Department of Pediatrics, Brody School of Medicine, East Carolina University; College of Nursing, East Carolina University; Department of Surgery, Brody School of Medicine, East Carolina University; Department of Pathology, Brody School of Medicine, East Carolina University; Department of Microbiology and Immunology, Brody School of Medicine, East Carolina University

**Keywords:** Seroprevalence, Young Adults, Longitudinal, Nucleocapsid Protein, Severe Acute Respiratory Syndrome Coronavirus 2 (SARS-CoV-2)

## Abstract

**Objective:** We conducted a longitudinal study to estimate immunity produced in response to severe acute respiratory syndrome coronavirus 2 (SARS-CoV-2) infection among university students over seven months.

**Methods:** All participants were attending a public university and resided in Pitt County, North Carolina. University students enrolled weekly for 10 weeks between August 26, 2020 and October 28, 2020, resulting in 136 young adults completing at least one study visit by November 17, 2020. Enrolled students completed an online survey and nasal swab collection at two week intervals and monthly blood collection between August 26, 2020 and March 31, 2021.

**Results:** Amongst 695 serum samples tested during follow-up, the prevalence of a positive result for anti-nucleocapsid antibodies (N-IgG) was 9.78%. In 22 students with more than one positive N-IgG serum sample, 68.1% of group had decline of N-IgG below positive threshold over 140 days. Anti-spike antibodies were detected in all 11 vaccinated students who were vaccinated during March 2021.

**Conclusions:** In healthy young adults, N-IgG wanes below detectable threshold within five months. S-IgG remained consistently elevated months after infection, and significantly increased after vaccination.

## Introduction

Severe Acute Respiratory Syndrome Coronavirus-2 (SARS-CoV-2), which causes coronavirus disease 19 (COVID-19), emerged in China in late 2019.^1^ SARS-CoV-2 belongs to the *Betacoronavirus* genus in the family *Coronaviridae*. The viron particle contains four primary structural virion proteins: nucleocapsid (N), spike (S), envelope (E), and membrane (M).

As young adults returned to university campuses in fall 2020, most institutions of higher education offered COVID-19 testing to those with symptoms and to contacts of confirmed or suspected cases.^2,3^ Testing asymptomatic persons in a congregate setting has been part of a comprehensive strategy to reduce transmission,^4,5^ since young adults may spread the infection while asymptomatic.^6-8^

Besides testing for active infection, one way to monitor virus prevalence and spread in a population is to measure specific antibodies to SARS-CoV-2 in sera. The common target antigens for serological assays are the nucleocapsid (N) protein and the spike (S) protein. The presence of anti-SARS-CoV-2 immunoglobin IgG (IgG) indicates the individual has been infected and mounted an immune response to the virus from current or prior infection. Among PCR positives, seroprevalence ranges from 88-100% in large studies.^9,10,11^ A vast majority of SARS-CoV-2 infected individuals seroconvert for a duration of months,^10^ and natural immunity may persist for longer than 12 months.^12^ In persons with asymptomatic or mild cases, IgG seroconversion takes a longer time to mount and the peak antibody response is lower than those with more serious systemic disease.^13-15^ IgG antibody responses wane over time,^9,16-18^ particularly Nucleocapsid antibodies.^15,19^ Importantly, the strength and longevity of the antibody response informs whether persons are likely to be protected from reinfection.^10,20,21^

Longitudinal studies have been conducted to examine the persistence of IgG antibody duration over time, mostly in healthcare workers^18,22,23^, long-term care residents^19^, and COVID-19 patients.^9,10,13,24-26^ Limited longitudinal evidence exists on seroprevalence conversion and reversion among healthy young adults enrolled at university. Reports suggest that peak antibody levels are lower in those with asymptomatic to mild infection,^9,16,15^ and rapid decline of IgG immunity has been documented in those who have recovered from COVID-19.^9,16,21,25^ Less is understood about immunity persistence in healthy young adults.

As part of a surveillance research program among a cohort of young adult university students, nucleocapsid (N) IgG (N-IgG) and spike (S) IgG (S-IgG) antibodies were measured. We hypothesized that IgG status would decline over time, consistent with data from healthcare workers. Herein, we report the seroprevalence of N-IgG, the time to N-IgG development, the occurrence of sero-reversion for N-IgG and compared this with the development of antibodies to Spike protein (S-IgG) among a group of university students over seven months.

## Materials and Methods

### Participant recruitment and eligibility

University and Medical Center Institutional Review Board (UMCIRB) approved th research (#20-002665). As part of reopening a large public university in North Carolina (East Carolina University, ECU, Greenville, NC, USA) during fall semester 2020, a surveillance research project was implemented that included testing a group of students bi-monthly for presence of active SARS-CoV-2 infection and monthly for development of humoral immunity. In spring semester 2021, this same cohort was invited once per month for three consecutive months to test for active infection and immunity. An undergraduate or graduate student was eligible if enrolled full or parttime in fall semester 2020, listed a Pitt County residential address, was 18 years of age or older, and spoke English. Students affiliated with the ECU athletics program were tested under a separate protocol and excluded.

### Sampling and Recruitment

Three sets of 300 students (50% Freshman) were randomly generated and the first set of 300 were contacted via email to participate. Due to multiple large clusters of COVID-19 disease among the student body, the dormitories were closed to most students beginning August 26, 2020 resulting in many randomly selected students returning to their home residence outside Pitt county. At that time, the additional 600 randomly selected students received an email link to reply with a phone number for study staff to screen for eligibility. Additional students were recruited during September and October through announcements posted on ECU social media pages and distributed to student organizations and departmental rosters (e.g. dance, theater, art). During fall semester 2020, students were enrolled weekly between August 20, 2020 until October 28, 2020 (Wave 1). Any participant with at least one study visit during Wave 1 was invited to continue during spring semester 2021 to complete a study visit on January 27, February 24, and March 31, 2021 (Wave 2).

### Data Collection

An online survey was required every two weeks concurrent with each clinic visit during fall 2020, and once monthly during spring 2021. A $20 incentive was provided when the survey and nasopharygeal (NP) swab collection were completed. The survey was updated to collect information on vaccination intent and completion of vaccination, respectively. Study data were collected and managed using REDCap (Research Electronic Data Capture), a secure, web-based software platform, hosted at East Carolina University.^27^

### Study visit procedures

Students received a date and time for the clinic visit every two weeks during fall 2020 and once monthly during spring 2021. Signed informed consent was obtained at the initial clinic visit for Wave 1 and Wave 2, respectively. Participant’s temperature and COVID-19 symptoms were recorded at check-in. There were 13 study visits in Wave 1 and 3 study visits in Wave 2. If a student missed a scheduled visit, he/she was requested to attend the next week’s clinic visit.

#### Nasopharygeal swab (NP)

A healthcare worker inserted a sterile swab approximately 2.5 inches in one preferred nostril and rotated swab five times clockwise and five times counterclockwise before removing. The swab was placed head down into viral transport tube and placed into a rack over ice packs in a portable cooler for approximately three hours before transport to the testing laboratory.

#### Serology

A trained phlebotomist collected approximately 5mL of venous blood into a serum separator tube using a butterfly needle for analysis of N-IgG. A second tube of blood was collected on a subgroup of 28 individuals re-consented during spring semester 2021 to identify antibodies to spike protein (S-IgG). The 28 students were: (1) 25 participants who were seropositive for N-IgG during fall 2020 or spring 2021; and (2) three participants who tested PCR positive as of November 17, 2020 but were seronegative for N-IgG on November 17, 2020. No samples were collected between November 18, 2020 and January 23, 2021. November 17, 2020 was as last study visit during fall semester 2020.

### Laboratory methods

#### RT-PCR

Active infection was identified from a positive RT-PCR assay from the NP sample. Vidant Medical Center Clinical Laboratory (VMCCL) conducted SARS-CoV-2 identification using the Lyra® Direct SARS-CoV-2 Assay, which is a real-time RT-PCR assay intended for the qualitative detection of RNA nucleic acid from SARS-CoV-2 in NP samples.

#### Antibodies

Laboratory analysis of immunoglobulin G (IgG) antibodies to the SARS-CoV-2 virus was conducted using a chemiluminescent microparticle immunoassay (CMIA) for the qualitative detection of IgG antibodies to the SARS-CoV-2 nucleoprotein (N-IgG). The recommended index value threshold of 1.4 signal-to-cutoff (S/C) ratio or above (≥1.4) was used to define IgG seroposivitity.^28,29^ We collected blood every four weeks to allow for IgG seroconversion, which ranges from 14 days to 4 weeks after symptom onset.^21,30^ Antibody titers were not performed on N-IgG blood samples.

Both IgG antibodies to the SARS-CoV-2 spike protein (S-IgG) and nucleocapsid protein (N-IgG) were examined at three time points: January 27, February 24, and March 31, 2021.

The spike protein, the structural protein often used as a target for characterizing the immune response to SARS-CoV-2, contains the receptor binding domain (RBD) that the virus uses to dock to its cellular receptor, antiogensin-converting enzyme-2.^13^

The S-IgG responses were measured using an enzyme-linked immunosorbent assay (ELISA) as per previous work.^31-33^ Ninety-six-well ELISA plates (Immulon H2B; Thermo Scientific) were coated with 0.09 µg/well of recombinant SARS-Cov-2 spike, (R&D System, Cat #10549-CV) in coating buffer (pH 9.8) at 4°C overnight. Plates were blocked with 2% fetal bovine serum in PBS at room temperature for 30 min. Plates were washed twice with ELISA wash buffer (1x PBS, 0.02% Tween 20, 0.1% NaN3), and human sera were added with a serial dilution of 1:3. Plates were incubated at room temperature for 2 hours and washed 3 times with ELISA wash buffer. Alkaline phosphatase (AP)-conjugated goat anti-human IgG (H+L) (1:2200, Promega) was added and incubated at room temperature for 2 h. Plates were washed 3 times and developed (alkaline phosphate substrate kit; Bio-Rad), and the absorbance was read at 405 nm in AccuSkan FC (Skanlt 6.1, Fisher Scientific). A positive result was defined as four-fold increase in optical density absorbance compared to the negative control value (0.06), or 0.24 optical density absorbance. The negative value was set based on result from two participants who tested PCR positive but did not have a positive nucleocapsid IgG at anytime during follow-up. The spike protein assay was performed in the lab of Dr. Rachel Roper, PhD (Brody School of Medicine).

### Outcome measures

The primary endpoints were N-IgG persistence and N-IgG seroreversion. Persistence was defined as the number of days between the date of initial positive N-IgG result and the date of last positive N-IgG result when both N-IgG values were positive. Sero-reversion (loss of N-IgG) was defined as the number of days between initial positive N-IgG result and the date of first negative N-IgG.^34^ N-IgG posivitiy is defined as S/C ratio ≥ 1.4. Date of PCR positive test to date of initial N-IgG positive was included in results, since the initial date of N-IgG positive is used to calculate persistence.

### Statistical analysis

Categorical data are summarized by N-IgG status (positive versus not positive) by presenting the number of patients and percentage for each category. Groups were compared using the chi-square test. Missing demographic and baseline data were treated as missing and were not imputed.

Duration of N-IgG seropositivity was summarized by N-IgG persistence status by presenting the the mean, standard deviation (SD), median, with minimum and maximum. Time to loss of N-IgG persistence was analyzed using the Kaplan-Meier method. Participants with persistent N-IgG (no loss below S/C ratio 1.4) were right censored (coded zero).

The N-IgG profile for individual participatns who were N-IgG positive were graphically displayed using spaghetti plots. Plots were used as guides to assess change in the N-IgG assay over time. Analyses were performed using Excel (Windows 10), GraphPad Prism v9, and Stata 14. All tests were 2-tailed with *P*-value less than 0.05 as statistically significant.

## Results

In Wave 1, 81 randomly selected and 55 self-selected eligible students completed at least one clinic visit. Among 136 consented students in Wave 1, 86 completed all scheduled study visits (63.2%) during Wave 1. Ninety-seven participants from Wave 1 reconsented for Wave 2 (71.3%). Twelve students completed all scheduled study visits between August 26, 2020 and March 31 2021. There were 13 study visits in Wave 1 and 3 study visits in Wave 2.

**Figure 1** displays enrollment in fall 2020 and spring 2021. Sixteen participants were N-IgG positive during September-October 2020 and 12 were positive during January (n=7), February (n=2), and March (n=3), 2021.

**Figure 1.**
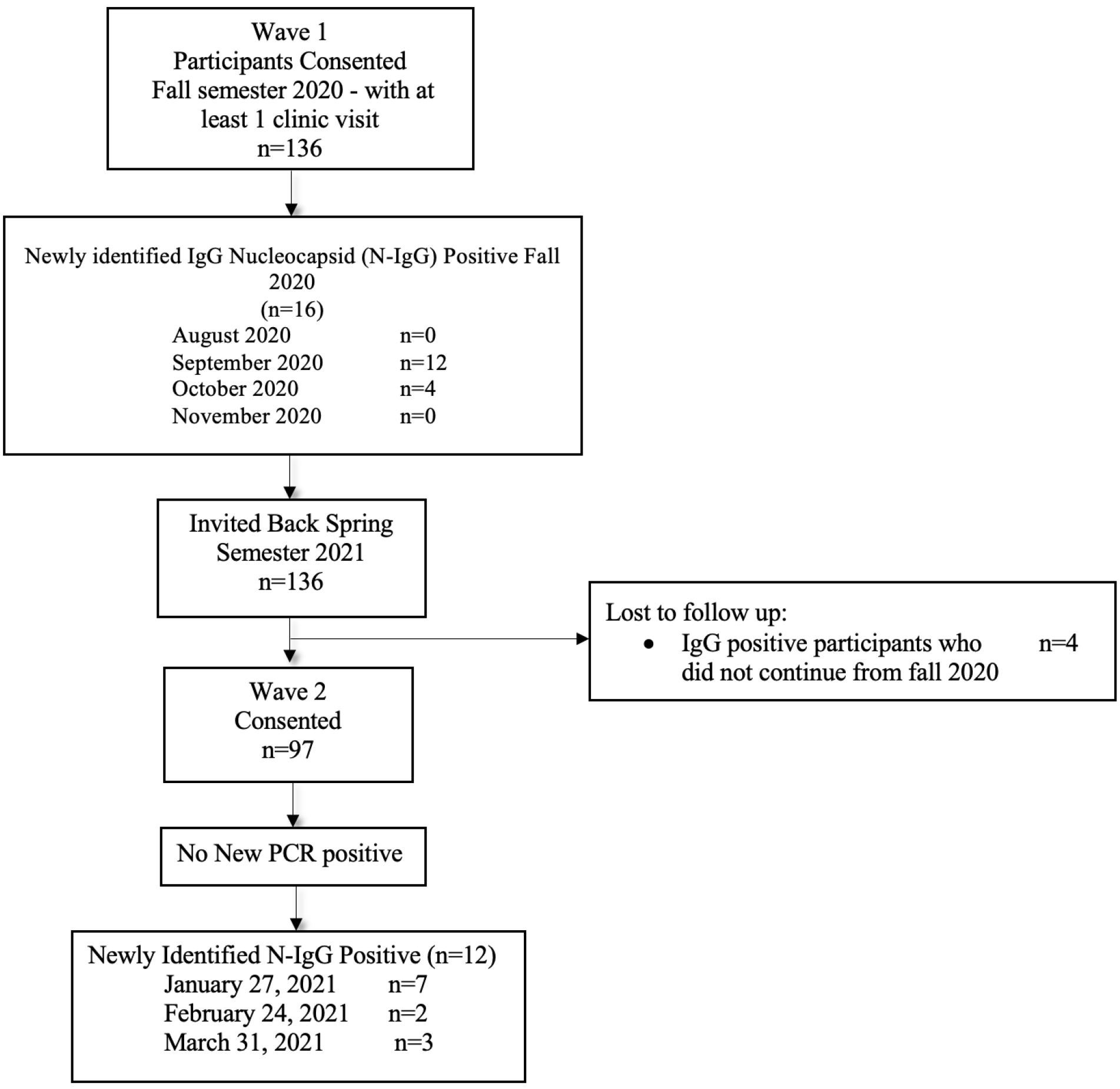
Participant Flow between Fall 2020 and Spring 2021

Ninety-four women (69%) and 42 men (31%) participated in at least one clinic visit during fall 2020. Seventy-one percent (71.43%) of N-IgG positive were ages 18 to 21 and 32% were freshman. A majority of N-IgG positive had obtained testing for COVID-19 prior to enrollment compared to N-IgG negative. No differences between groups were found for having a positive COVID-19 test or completing one or two doses of vaccine (**Table 1**).

**Table 1.** Characteristics of Study Population by Nucleocapsid IgG (N-IgG) Status

|  | Ever N-IgG Positive |  | Always N-IgG Negative |  |
| --- | --- | --- | --- | --- |
|  | Number | %* | Number | % |
| Age |  |  |  |  |
| 18 - 20 | 20 | 71.43 | 43 | 39.81 |
| 21- 22 | 6 | 21.43 | 45 | 41.67 |
| 23 -25 | 1 | 3.57 | 12 | 11.11 |
| 26+ | 1 | 3.57 | 8 | 7.41 |
| Total | 28 | 100 | 108 |  |
| Biologic sex |  |  |  |  |
| Male | 9 | 32.14 | 33 | 30.56 |
| Female | 19 | 67.86 | 75 | 69.44 |
| Total | 28 | 100 | 108 |  |
| Race |  |  |  |  |
| White/Caucasian | 21 | 75 | 86 | 78.7 |
| Black/African American | 3 | 10.71 | 8 | 7.41 |
| Asian | 1 | 3.57 | 7 | 6.48 |
| Other | 3 | 10.71 | 2 | 1.85 |
| Total | 28 | 100 | 6 | 5.56 |
| Ethnicity |  |  |  |  |
| Latinx | 6 | 22.22 | 10 | 9.43 |
| not Latinx | 21 | 77.78 | 96 | 90.57 |
| Total | 27 | 100 | 106 |  |
| School Year, fall 2020 |  |  |  |  |
| Freshman | 9 | 32.14 | 19 | 17.59 |
| Sophomore | 6 | 21.43 | 7 | 6.48 |
| Junior | 6 | 21.43 | 19 | 17.59 |
| Senior | 5 | 17.86 | 43 | 39.81 |
| Graduate School | 2 | 7.14 | 20 | 18.52 |
|  | 28 | 100 | 108 |  |
| Had a COVID-19 Test before enrollment |  |  |  |  |
| No | 6 | 21.43 | 52 | 48.15 |
| Yes | 22 | 78.57 | 55 | 50.93 |
| Do not know | 0 | 0 | 1 | 0.93 |
| Total | 28 | 100 |  |  |
| COVID-19 Vaccine as of 3/31/21** |  |  |  |  |
| First dose | 11 | 39.3 | 28 | 40.58 |
| Second dose | 3 | 10.71 | 17 | 24.64 |
| No dose of vaccine | 14 | 50 | 24 | 34.78 |
| Total | 28 | 100 | 69 | 100 |
| Tested COVID-19 positive before enrollment |  |  |  |  |
| Positive | 8 | 28.57 | 2 | 0.02 |
| Negative | 13 | 46.43 | 52 | 48.15 |
| Waiting on result | 0 | 0.00 | 1 | 0.01 |
| Inconclusive | 1 | 0.04 | 0 | 0.00 |
| missing | 6 | 21.43 | 53 | 49.10 |
|  | 28 |  | 108 |  |
| Flu Vaccine 2019-2020 season |  |  |  |  |
| No | 14 | 50 | 36 | 33.33 |
| Yes | 11 | 39.29 | 65 | 60.19 |
| Do not know | 3 | 10.71 | 7 | 6.48 |
|  | 28 |  | 105 |  |

\*Some percentages do not add to 100 due to missing values.
\*\* Denominator is 97; vaccine offered in spring semester 2021

### SARS-CoV-2 PCR results

A new COVID-19 case was defined as a positive PCR test without concurrently testing positive for SARS-CoV-2 antibodies. Four women met the case definition for positive PCR test (2.94%) at the Wave 1 visit. The two women that tested PCR positive at the study visit and reported symptoms (loss of taste and smell, chills with muscle aches, respectively) mounted a positive (≥1.4 S/C) N-IgG antibody response. In two asymptomatic women who tested PCR positive at the study visit, N-IgG antibodies were not detected (never above 0.01 S/C). It is possible that two asymptomatic women had false positive PCR results or they had transient superficial infections that did not result in substantial systemic antibody responses, as has been reported by others.^14^

During Wave 2, two COVID-19 cases were detected at the monthly clinic visit. These two individuals were also N-IgG positive on the same day as PCR testing. Based on the COVID-19 case definition used by the State of North Carolina during Spring 2021, the positive PCR tests were not counted as a newly identified COVID-19 cases. Among 32 students that had evidence of past COVID-19 infection based on N-IgG, 15 were self-reported PCR positive cases.

### Persistence and loss of anti-nucleocapsid antibodies

The range of N-IgG values was 0.01 to 7.01. At the time of blood collection, 28 participants tested N-IgG positive (S/C ≥ 1.4) during the study interval (**Figure 2**). The proportion of newly identified positive N-IgG was 11.76% (16/136) between August 26, 2020 and November 17, 2020. Between January 27 and March 31, 2021, the prevalence for newly identified N-IgG was 12.37% (12/97). Amongst all serum samples tested, 9.78% (68/695 [627+68=695]) detected N-IgG positive between August 26, 2020 and March 31, 2021.

**Figure 2.**
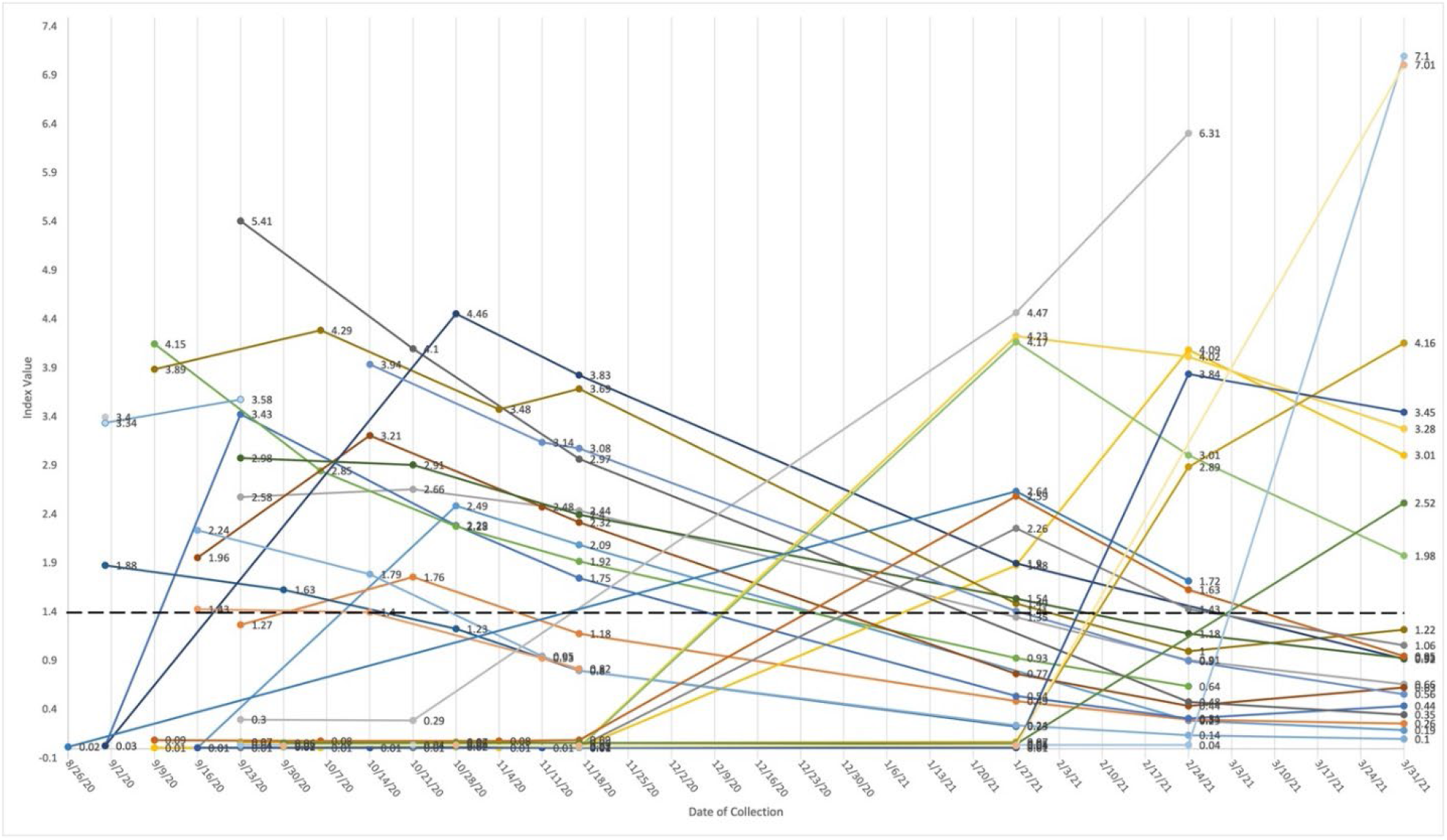
Change in quantity of anti N-IgG for 28 young adults with at least one N-IgG positive result, August 26, 2020 – March 31, 2021 Each individual is represented by a colored line from first testing date to last testing date. Number at dot is index value. An index value greater than or equal to (≥) 1.4 is positive (bold dotted line). The chemiluminescent microparticle immunoassay (CMIA) captures antibodies reactive with the Nucleocapsid (N) protein and specifically detects IgG only. This qualitative assay reports a ratio of luminescence between sample and calibrator (the S/CO index) (Abbott Architect i2000 2). One N-IgG positive participant enrolled and withdrew in September (2 data points); one participant had one study visit in September 2020 (1 data point). Three participants tested positive at last study visit date (1 data point). The remaining 23 provided N-IgG measures across multiple weeks.

At end of Wave 1, 43.5% (10/23) of N-IgG measures had declined below the positivity value (S/C ≥1.4). Seven new N-IgG positive tests were reported on January 27, 2021, and of those, four lost positive N-IgG status by the end of Wave 2 (March 31, 2021) (57.1%). Three tested N-IgG positive for the first time on March 31, 2021, the last scheduled blood collection day.

Over 16 study visits, the percentage of positive N-IgG among those scheduled to provide a blood sample ranged from 0.45% to 14.3%. The mean N-IgG levels among any positives ranged from 1.88 to 4.06 (**Figure 3**).

**Figure 3.**
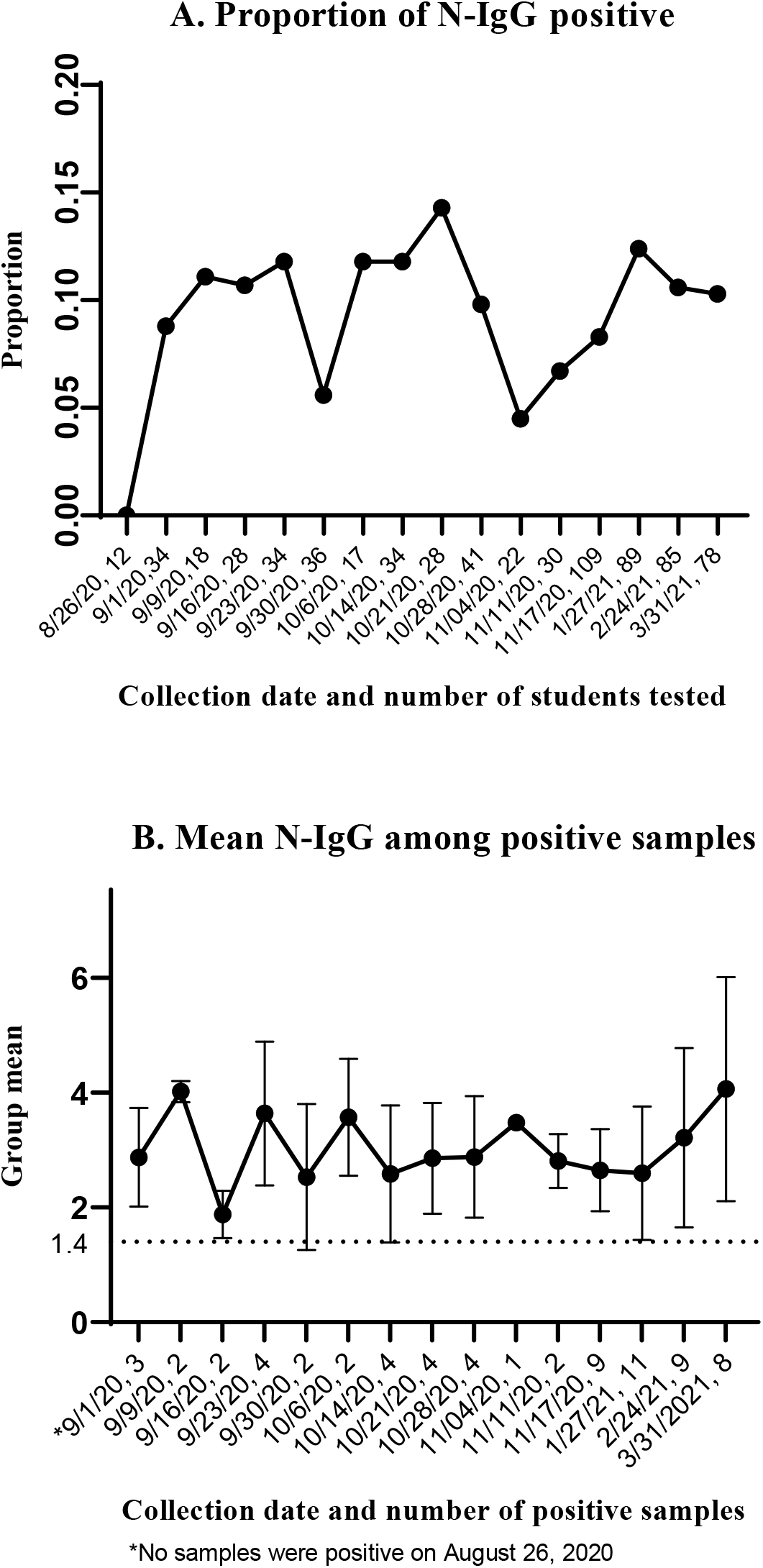
Descriptive Statistics of nucleocapsid (N) IgG among young adults by collection date

Twenty-three of 28 students had more than one positive N-IgG result. Persistent N-IgG positivity and loss were estimated on these 23 students (**Table 2**). Sixty-five percent (15/23) of participants with more than one positive N-IgG test lost seropositivity N-IgG status. The mean number of days between a positive PCR test and the first measure of positive N-IgG antibodies was 21.21 (14.76, SD) days. The mean and median duration of N-IgG persistence was 54.3 and 48 days, respectively (defined as the number of days between the first positive N-IgG result and the last positive N-IgG result, when both measures remained positive).^34^

**Table 2.** Number of days of persistence and loss for N-IgG seropositivity in 23 young adults with greater than one positive N-IgG result, September 1, 2020 through March 31, 2021*a*

| Number of Days | PCR positive test to Initial N-IgG Positive (n=19) <sup>b</sup> | N-IgG persistence n=23 <sup>d</sup> | Loss of Persistence <sup>e</sup> n=15 <sup>f</sup> |
| --- | --- | --- | --- |
| Mean, SD | 21.21 (14.76) | 54.30 (33.51) | 82.33 (44.16) |
| Median | 19 | 48 | 63 |
| Range | 0-54 <sup>c</sup> | 20-140 | 28-154 |

SD, standard deviation
<sup>a</sup> Serology was offered approximately every 30 days from date of first collection, excluding December 2020.
<sup>b</sup> Three participants were missing date of positive PCR test. One participant who self-reported PCR positive and tested IgG positive in September 2020 returned on January 27, 2021. This outlier was removed from Table 2 leaving 19 PCR positive students; column 2 with participant's values (days) were: Mean, SD: 27.55 (31.78), Median: 19.5, and Range: 0-148 days.
<sup>c</sup> Two tested PCR positive and IgG positive on same day; the number of days between PCR positive and IgG positive was zero.
<sup>d</sup> IgG persistence is defined as number of days between date of initial positive N-IgG and date of last positive N-IgG.
<sup>e</sup> Loss of IgG (days from initial IgG positive to first negative IgG result (S/C ratio <1.4).
<sup>f</sup> Excludes 8 with no decline of persistence.

**Figure 4** displays the number of days from initial N-IgG positive to last measure of N-IgG positive (column 3 from Table 2, ‘IgG persistence’) for 22 students. At 140 days (∼4.7 months), 68.1% (15/22) of participants had lost positive N-IgG status (below 1.4 S/C threshold). The mean (SD) and median number of days were 77.8 (9.9) and 62, respectively, with interquartile estimates between 29 days to 105 days. One participant was lost to follow-up in September 2020 and excluded from **Figure 4** (but remained in Table 2).

**Figure 4.**
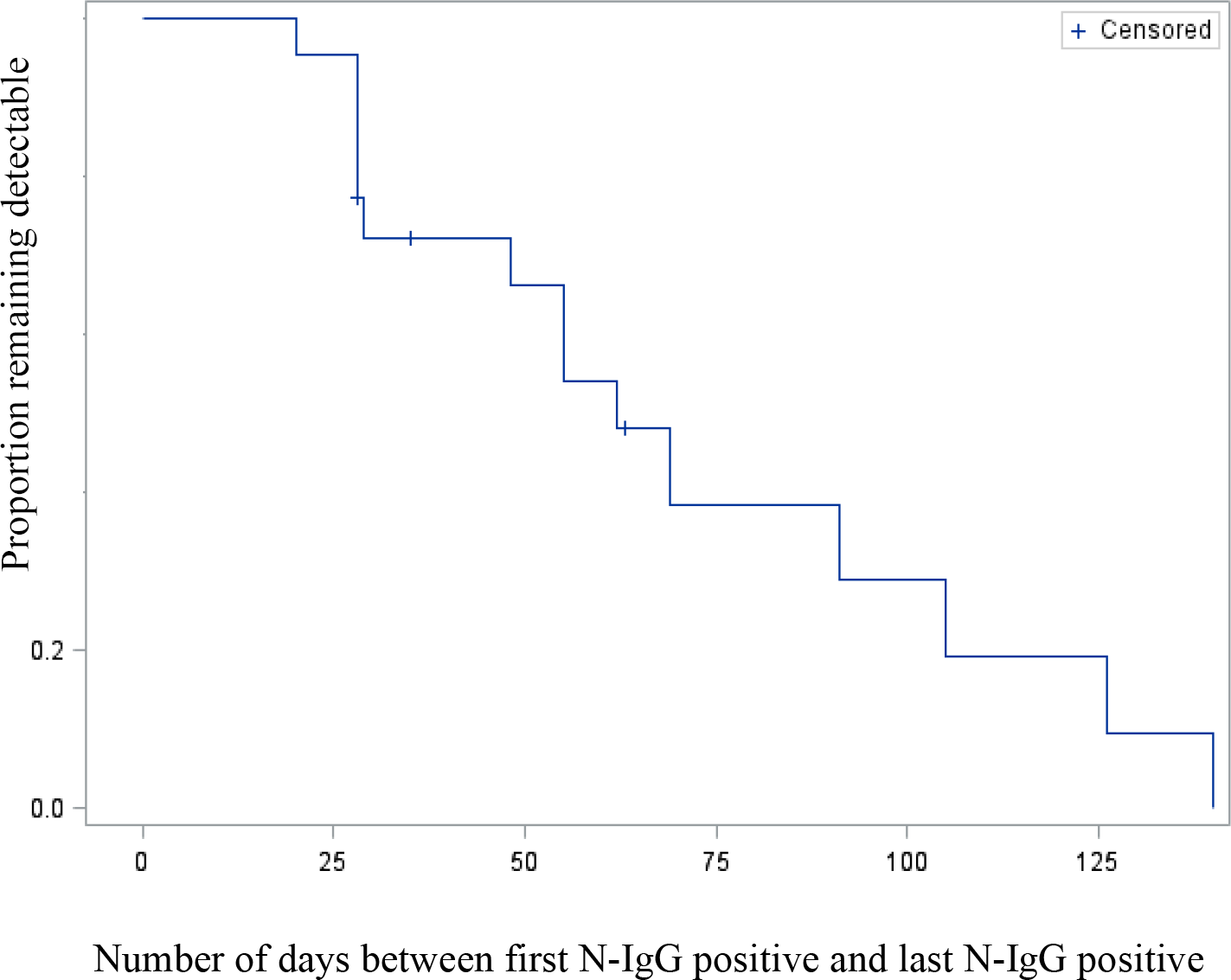
Decline in nucleocapsid IgG among 22 young adults Decline in positive N-IgG status over 140 days (n=22). One observation in table 2, column 3 was excluded from Figure 3 due to withdrawal from participation in September 2020. X-axis is days between 0 to 140 days of N-IgG persistence by end of follow-up. Y-axis is proportion of people remaining with IgG persistence. Seven (censored) had persistent detectable N-IgG. Persistence is defined as number of days between first positive and last positive IgG nucleocapsid result. This graph does not reflect number of days between date of first positive PCR test positive and date of first IgG positive. 68.1% had decline of N-IgG over 140 days.

### Presence of anti-spike antibodies

Among 56 samples (25 students), 91% (51/56) of serum samples had detectable anti spike IgG (S-IgG) antibodies (**Figure 5**) at any of three time points (January 27, February 24, or March 31). There was no significant difference in S-IgG levels for 9 students who were tested on both January 27 and February 24, 2021 (p=0.23). For nine students who were tested on both February 24 and March 31, 2021, no significant difference in S-IgG levels were found (p=0.07). However among nine vaccinated students (at least 1 dose) on March 31, 2021, a significant difference in S-IgG was found between February 24 and March 31, 2021 (p=0.02).

**Figure 5.**
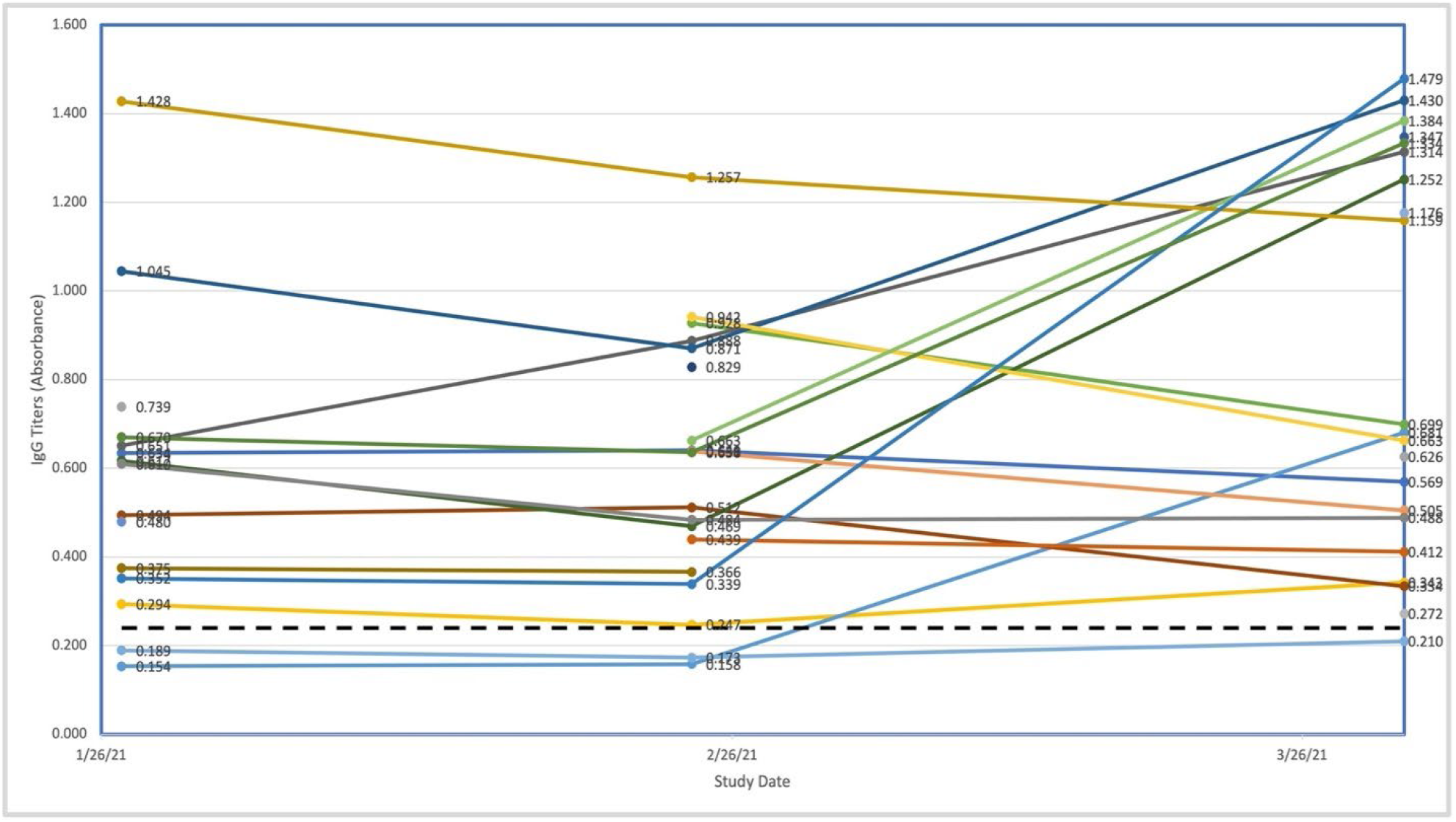
Change in anti-Spike Protein-IgG for 25 young adults over January, February, and March 2021 by vaccination status Anti Spike-IgG was analyzed using sera collected on January 27, 2021, February 24, 2021, and March 31, 2021. Each color is a unique person. X-axis is date of collection for 25 young adults. Y-axis is ELISA absorbance to anti S-IgG. A positive result was defined as four-fold the negative control value (0.06), or 0.24 optical density absorbance. On March 31, 2021, vaccinated participants had the following S-IgG values, dose-vaccination date: 0.343, Dose 1- 3/27/21; .0681, Dose 2- 3/30/21; 0.334, Dose 1- 3/31/21; 1.314, Dose 1- 3/20/21; 1.430, Dose 1- 3/19/21; 1.252, Dose 1- 3/25/21; 1.384, Dose 1- 3/11/21; 1.347, Dose 1&2- 3/3/21 & 3/30/21; 1.479, Dose 1- 3/5/21; 1.334, Dose 1- 3/14/21; 1.176, Dose 1&2- 1/27/21 & 2/17/21.

On January 27, 2021 (the first study visit during spring semester 2021), six of 12 (50%) participants tested for both S-IgG and N-IgG were S-IgG positive and N-IgG negative (discordant). Among 14 participants with both N-IgG and S-IgG analyzed on March 31, 2021, 14 had detectable S-IgG (100%) and 9 had negative N-IgG (64.2%) (discordant). Among four participants with positive N-IgG, three tested antibody positive for the first time on the day of sample collection. One of 13 S-IgG measures was slightly above the 0.24 S-IgG threshold (0.272). All 11 participants vaccinated during March 2021 had detectable anti S-IgG on March 31, 2021.

Two participants, who tested PCR positive in fall 2020, never had detectable N-IgG (never >0.01) and did not have detectable S-IgG during January or February. Participant Q, who was not vaccinated, remained negative for detetable N-IgG and S-IgG in March 2021. Participant E was vaccinated on March 30; S-IgG between February 27, 2021 and March 31, 2021 changed by 331% for participant E, but N-IgG was not detected.

## Discussion

Repeated measures of serum antibody in a young adult university cohort serves to inform the capacity for adaptive immunity through antibody persistence and loss. Among a group of 22 young adults who were followed over seven months, we found that N-IgG positive reverted to below threshold in 68.1% of participants over 140 days. Levels of S-IgG were markedly elevated in those recently vaccinated beginning in January 2021. A decline in S-IgG also occurred over thee months but samples absorbance positive in January 2021 remained S-IgG positive on March 31, 2021 in the absence of vaccination. Our results are consistent with findings that the anti S-IgG confers more persistence than anti N-IgG.^10,15,19^ This may be due to the initial response to S-IgG is stronger due to its fundamental immunogenicity, the large quantity of S-IgG produced during infection, or the fact that it is expressed both on the virus particle and on the surface of infected cells. This is in contrast to N-IgG, which is sequestered inside the virus particle or inside the infected cell. Alternatively, the laboratory specificity of anti-spike antibodies may be higher than for anti nucleocapsid antibodies. This specificity cannot be directly compared, because the S and N proteins are fundamentally and biochemically different proteins and different reagents are used to detect them.

Longitudinal studies conducted in a non-hospitalized young adult population that examined immunoglobulin seroconversion and seroreversion were not identified. Several studies have examined seroconversion and loss over five months or more in clinic patients. In a group of COVID-19 patients, Maine and colleagues tested 427 sequential COVID-19 serum samples between March and August 2020 collected up to 168 days post onset of symptoms.^25^ The median days from IgG seronegative to seroconversion was 11.5 days, and 10 patients followed greater than 100 days all experienced IgG decline, similar to our findings.^25^ Among 45 Belgian COVID-19 cases with mild disease (defined as asymptomatic or not needing hospitalization), 61.1% were seronegative within 6 months after first PCR positive.^14^ In two outbreaks at the same long term care facility, all ten residents in both outbreaks has a significant anti-N antibody decline, with all ten second serology measures remaining barely positive after seven months; decline in anti-spike over four months was not significantly different, similar to our findings.^19^

Among cohorts of healthcare workers, similar declines in N-IgG were noted. Anna and colleagues found anti-N titers declined by 31% over 4-8 weeks in the majority of French healthcare workers.^18^ In cohort of hospital staff in Wales, four months after a positive PCR test, only 22% (2/9) had detectable antibodies against nucleocapsid protein, while 78% (7/9) individuals had detectable antibodies against spike protein.^23^ Given that 65% of students lost detectable N-IgG over 140 days, our findings are consistent with other studies in non-hospitalized adults. 46% of our study population were between ages 18-20. Our data suggest that young adults who have asymotmatic or mild disease have a rapid peak and more rapid decline in peak N-IgG antibody response than hosptialized patients.

Strengths of this study include the longitudinal design in a healthy young adult university-based population. Compliance with scheduled study visits was high in both study segments (fall semester 2020 and spring semester 2021). Limitations include the absence of both S-IgG and N-IgG titers during wave 1. Gaps in monthly blood collection (approximately every 30 days) due to some missed appointments may dilute important changes over time. Thirty days between each sera collection may inflate the number of days between persistence or seroreversion. Since the study population was students currently enrolled at our university, we decided against storing sera for future analysis. We did not collect symptoms or disease severity for self-reported COVID-19 disease that occurred prior to enrollment. We did not collect a second tube on all 97 consented students in spring 2021 due to budgetary constraints and have not collected additional data on re-infection or other measures since March 2021 for the same reasons. This student population participating in this study may not be representative of the ECU student population as a whole or young adults in general. Seropositivity was assessed in students who were permitted to attend in-person courses on campus, and/or live in the dorms, and otherwise be motivated to travel to the campus-based testing clinic for a bi-monthly visit. This study was implemented six months after the pandemic was declared in March 2020. The alpha, beta, and gamma variants were the dominant strains in NC during Wave 1 and Wave 2. As the pandemic has progressed, greater understanding about IgG serology assays has emerged along with significant expansion of laboratory capabilities. Reliance on detection of N-IgG antibodies exclusively for determining potential COVID-19 immunity should be used with caution and multiple independent assays may improve accuracy of estimating seroprevalence over time.^15,35^

In conclusion, this study adds to understanding that healthy young adults mount an antibody response that waned in 68% of participants over 140 days. The S-IgG response declined non-significant over 30 days in the absence of vaccination and rose sharply after vaccination.

## Data Availability

All data produced in the present study are available upon reasonable request to the first author.

## Acknowledgements

The authors wish to thank Drs Silvana Pasetto and Ming Fan and Evan Bradner for their help collecting samples and technical expertise. We appreciate the dedication of ECU students who contributed to this research.

## Data Availability

Data availability can be made through contacting corresponding author.

